# A machine learning approach for identification of gastrointestinal predictors for the risk of COVID-19 related hospitalization

**DOI:** 10.1101/2021.08.27.21262728

**Authors:** Peter Lipták, Peter Banovčin, Róbert Rosoľanka, Michal Prokopič, Ivan Kocan, Ivana Žiačikova, Peter Uhrík, Marian Grendár, Rudolf Hyrdel

**Affiliations:** Gastroenterology Clinic, University Hospital in Martin, Jessenius Faculty of Medicine in Martin (JFM CU), Comenius University in Bratislava, Slovakia; Clinic of Infectology and Travel Medicine, University Hospital in Martin, Jessenius Faculty of Medicine in Martin (JFM CU), Comenius University in Bratislava, Slovakia; Clinic of Pneumology and Phthisiology, University Hospital in Martin, Jessenius Faculty of Medicine in Martin (JFM CU), Comenius University in Bratislava, Slovakia; Laboratory of bioinformatics and biostatistics, Biomedical Centre Martin, Jessenius Faculty of Medicine, Comenius University, Mala Hora 4C, 036 01 Martin, Slovakia; Laboratory of theoretical methods, Institute of Measurement Science, Slovak Academy of Sciences, Dubravska cesta 9, 841 04 Bratislava, Slovakia

**Keywords:** COVID-19, SARS-CoV-2, Machine learning, Artificial intelligence, Random forest, Symptoms, Liver, Predictors, Hospitalization

## Abstract

**Background and aim:** COVID-19 can be presented with various gastrointestinal symptoms. Shortly after the pandemic outbreak several machine learning algorithms have been implemented to assess new diagnostic and therapeutic methods for this disease. Aim of this study is to assess gastrointestinal and liver related predictive factors for SARS-CoV-2 associated risk of hospitalization.

**Methods:** Data collection was based on questionnaire from the COVID-19 outpatient test center and from the emergency department at the University hospital in combination with data from internal hospital information system and from the mobile application used for telemedicine follow-up of patients. For statistical analysis SARS-CoV-2 negative patients were considered as controls to three different SARS-CoV-2 positive patient groups (divided based on severity of the disease).

**Results:** Total of 710 patients were enrolled in the study. Presence of diarrhea and nausea was significantly higher in emergency department group than in the COVID-19 outpatient test center. Among liver enzymes only aspartate transaminase (AST) has been significantly elevated in the hospitalized group compared to patients discharged home. Based on random forest algorithm, AST has been identified as the most important predictor followed by age or diabetes mellitus. Diarrhea and bloating have also predictive importance although much lower than AST.

**Conclusion:** SARS-CoV-2 positivity is connected with isolated AST elevation and the level is linked with the severity of the disease. Furthermore, using machine learning random forest algorithm, we have identified elevated AST as the most important predictor for COVID-19 related hospitalizations.

## Introduction

Acute SARS-CoV-2 infection presents with variable symptoms associated with various organ systems. Typical symptoms of COVID-19 are fever, cough, and in the case of a more severe course, dyspnea with respiratory insufficiency occurs [1]. In addition, COVID-19 may present with gastrointestinal symptoms, which include dominantly nausea, vomiting, diarrhea, anorexia and abdominal pain with relatively wide range of prevalence among different published studies [2]–[8]. Since COVID-19 pandemic is the cause of immense world health crisis, new diagnostic and therapeutic methods are rapidly emerging [9]. The use of artificial intelligence is just one of them. Shortly after the COVID-19 outbreak various machine learning algorithms have been implemented [10]–[13]. Machine learning helps to quickly identify patterns and trends of the large volume of data, that are difficult for humans to recognize [14]. The availability of objective stratification tools to rapidly assess a patient status and prognosis is of a great use for the frontline health-care providers [15].

The primary aim of this study is to assess possible predictive factors for SARS-CoV-2 outcome based on gastrointestinal symptoms and liver related laboratory results using machine learning algorithms of random forest [16], [17]. The secondary aim is to determinate the prevalence of gastrointestinal symptoms among patients with COVID-19 within different groups based on the severity of the disease.

## Methods

The Study had been performed from February through May 2021. Only persons of 18 years or older were included in the study. All patients enrolled in this study signed the informed consent. This study was approved by the Ethical committee of the University hospital in Martin, decision number: 14/2021.

2 distinct kinds of population had been considered for this study. First group consist of people who underwent nasopharyngeal swab in the outpatient hospital testing center for COVID-19 in order to determine whether they were SARS-CoV-2 positive. The method of SARS-CoV-2 detection from nasopharyngeal swab was PCR (polymerase chain reaction). This group was then subdivided based on their positivity. The negative group was thereafter used as a control group for this study. Second group consist of patients who attended COVID-19 emergency department (ED) in the University hospital. These patients were confirmed positive from nasopharyngeal swab either by PCR or antigen method. Only patients with typical COVID-19 symptoms (fever, cough, dyspnoe) were included in this study. Patients who were SARS-CoV-2 positive but didn’t present with typical COVID-19 symptoms (e.g. patients who came to emergency room because of other diagnoses but simultaneously were SARS-CoV-2 positive) were excluded. Therefore, we considered for this study only patients who were both tested positive and have at least one typical COVID-19 symptom.

This second group was then divided based on further evaluation and course of the disease. First subgroup consists of patients that didn’t require admission to the hospital and were referred to the outpatient care. Second subgroup of patients was admitted to the hospital. Consequently, this group was observed until end of hospitalization either because of death or resolution of the disease. This subgroup was also divided for analysis purposes between patients who required only standard hospital care and those who needed intensive care unit (ICU).

Data collection was based on questionnaire in the group from COVID-19 outpatient test center at the University hospital. Data from emergency room were obtained from the same questionnaire which was combined with information from medical examination by attending physician and from the mobile application MEDAsistent used for telemedicine follow-up developed at the Clinic of Pneumology and Phthisiology in the University Hospital in Martin. Further information (including laboratory results, chest X-ray etc.) about patients who were hospitalized was obtained from hospital information system.

The questionnaire consists of questions related to the present health complaints typical for COVID-19 and the spectrum of most common gastrointestinal symptoms which had occurred in the last 5-7 days before examination. Patients were also allowed to write down other presented symptoms in the case they were not in the original list. To include only new or worsened gastrointestinal symptoms in the study the questionnaire also included questions about chronic gastrointestinal symptoms and their possible worsening in the last 5-7 days before examination.

### Data analysis

The data were visualized and analyzed in R [18], ver. 4.0.5, with the aid of libraries gtsummary [19], rstatix [20], DescTools [21], randomForestSRC [22], PCAmixdata [23] and ggpubr [24]. The sample median and the lower and upper quartiles were used to summarize the data on continuous variables (e.g., age); counts and percentages were used to summarize factors (e.g., gender). The Chi-squared or Fisher test were applied to test the null hypothesis of independence between factors, followed, where appropriate, by the multiple comparisons with the Benjamini Hochberg adjustment. Using a contingency table, an absence of trend was tested by Cochran Armitage test. The null hypothesis of the equality of the population medians of a continuous variable was tested by the Kruskal Wallis test, followed by the Dunn multiple comparisons test with the Benjamini Hochberg correction of p-values. Two-way ANOVA was used to model the association between AST and group (Discharged home, Admitted to hospital) in interaction with recent ATB use and chronical liver disease (yes, no). The AST values were log-transformed to bring data to normality. Normality of residuals was assessed by the quantile-quantile plot with the 95% confidence band constructed by bootstrap. Assumption of homogenity of variance was tested by the Levene test. In order to assess predictive power of the gastrointestinal parameters and other measured variables for predicting outcome of the patient group the Random Forest Machine Learning algorithm was trained on the data. The predictive ability was quantified by the ROC curve, constructed from the Out-of-Bag data. Importance of the predictors was measured by the Variable Importance. A 2D representation of the data was obtained by Principal Component Analysis for mixed type of data. Findings with the p-value below 0.05 were considered statistically significant.

## Results

Total of 710 patients were enrolled in the study. 30 patients were excluded from the further analysis after primary screening. 352 participants from the outpatient center who were tested PCR negative for SARS-CoV-2 virus were considered as the control group. SARS-CoV-2 positive group from outpatient center had 166 participants. 162 patients from emergency department were enrolled. From this group 78 were discharged home, 57 admitted to hospital with standard care up until discharged from hospital. 27 patients required intensive care unit. Based on age, the groups from outpatient center had almost similar median of 42 and 41 years of age respectively. Hospitalized patients were significantly older as shown in the table 1. The presence of typical COVID-19 symptoms such as fever and cough were significantly higher in the hospitalized groups as opposed to outpatient participants. There were no significant differences based on sex.

**Table 1.**

|  |  | Outpatient<br>test center<br>(SARS-CoV-2 negative)<br>(n=352)<sup>1</sup> | Outpatient<br>test center<br>(SARS-CoV-2 positive)<br>(n=166)<sup>1</sup> | Discharged<br>home<br>(n=78)<sup>1</sup> | Admitted<br>to hospital-<br>standard<br>care<br>(n=57)<sup>1</sup> | Admitted<br>to hospital-ICU<br>(n=27)<sup>1</sup> | p value<sup>2</sup> |
| --- | --- | --- | --- | --- | --- | --- | --- |
| **Age** |  | 42 (31, 51) | 41 (32, 52) | 48 (42, 58) | 55 (46, 67) | 65 (55, 72) | <0.001 |
| **Gender** |  |  |  |  |  |  | 0.07 |
|  | female | 206 (59%) | 104 (63%) | 41 (53%) | 27 (47%) | 11 (41%) |  |
|  | male | 141 (41%) | 62 (37%) | 37 (47%) | 30 (53%) | 16 (59%) |  |
| **Fever** |  |  |  |  |  |  | <0.001 |
|  | Not-pre-<br>sented | 340 (97%) | 146 (88%) | 48 (62%) | 25 (44%) | 15 (56%) |  |
|  | Presented | 12 (3.4%) | 20 (12%) | 30 (38%) | 32 (56%) | 12 (44%) |  |
| **Cough** |  |  |  |  |  |  | <0.001 |
|  | Not-pre-<br>sented | 342 (97%) | 144 (87%) | 48 (62%) | 24 (42%) | 14 (52%) |  |
|  | Presented | 10 (2.8%) | 22 (13%) | 30 (38%) | 33 (58%) | 13 (48%) |  |
| **Diarrhea** |  |  |  |  |  |  | <0.001 |
|  | Not-pre-<br>sented | 330 (94%) | 141 (85%) | 64 (82%) | 34 (60%) | 19 (70%) |  |
|  | Presented | 22 (6.2%) | 25 (15%) | 14 (18%) | 23 (40%) | 8 (30%) |  |
| **Constipation** |  |  |  |  |  |  | 0.37 |
|  | Not-pre-<br>sented | 347 (99%) | 164 (99%) | 76 (97%) | 55 (96%) | 26 (96%) |  |
|  | Presented | 5 (1.4%) | 2 (1.2%) | 2 (2.6%) | 2 (3.5%) | 1 (3.7%) |  |
| **Bloating** |  |  |  |  |  |  | 0.13 |
|  | Not-pre-<br>sented | 341 (97%) | 161 (97%) | 76 (97%) | 51 (89%) | 26 (96%) |  |
|  | Presented | 11 (3.1%) | 5 (3.0%) | 2 (2.6%) | 6 (11%) | 1 (3.7%) |  |
| **Nausea** |  |  |  |  |  |  | <0.001 |
|  | Not-pre-<br>sented | 340 (97%) | 147 (89%) | 70 (90%) | 45 (79%) | 23 (85%) |  |
|  | Presented | 12 (3.4%) | 19 (11%) | 8 (10%) | 12 (21%) | 4 (15%) |  |
| **Heart burn** |  |  |  |  |  |  | 0.86 |
|  | Not-pre-<br>sented | 341 (97%) | 163 (98%) | 76 (97%) | 56 (98%) | 26 (96%) |  |
|  | Presented | 11 (3.1%) | 3 (1.8%) | 2 (2.6%) | 1 (1.8%) | 1 (3.7%) |  |
| **Abdominal pain** |  |  |  |  |  |  | 0.07 |
|  | Not-pre-<br>sented | 334 (95%) | 150 (90%) | 71 (91%) | 49 (86%) | 26 (96%) |  |
|  | Presented | 18 (5.1%) | 16 (9.6%) | 7 (9.0%) | 8 (14%) | 1 (3.7%) |  |

<sup>1</sup>Statistics presented: n (%); Median (IQR).
<sup>2</sup>Statistical tests performed: Kruskal-Wallis test; Fisher's Exact Test for Count Data with simulated p-value (based on 2000 replicates)

### Gastrointestinal symptoms occurrence and laboratory findings (table 1 and 2)

The presence of diarrhea, constipation, bloating, nausea, heartburn and abdominal pain was considered in this study. Presence of diarrhea and nausea was significantly higher in emergency department group than in the COVID-19 outpatient test center. Comparing SARS-Cov-2 negative and SARS-CoV-2 positive participants the presence of these symptoms has been more than three times higher in the positive group than in the negative one. This trend goes further considering ED patients and the severity of disease.

**Table 2.**
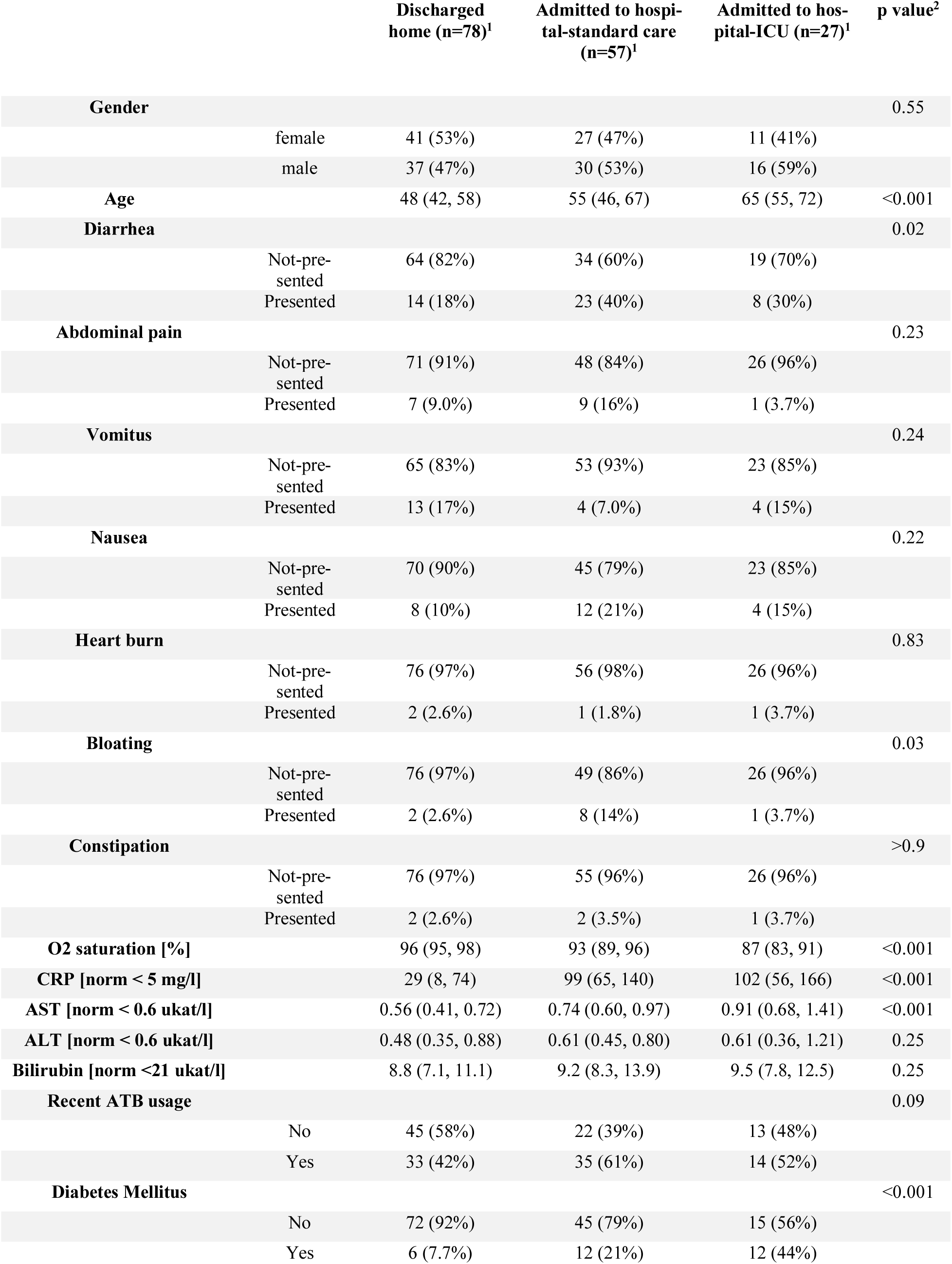
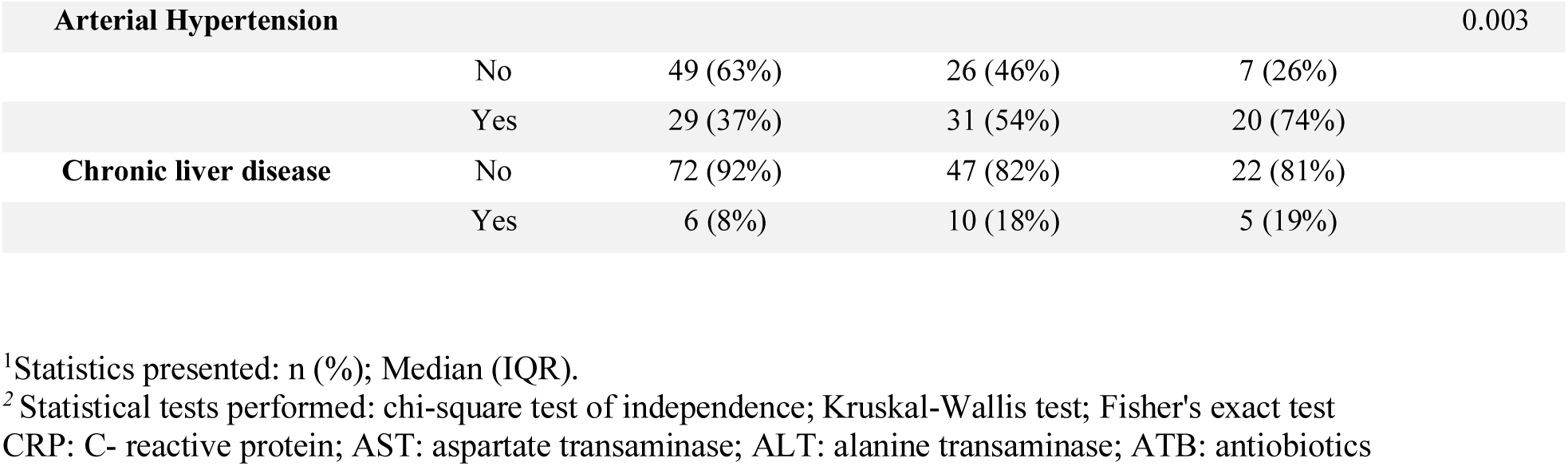

Among gastrointestinal symptoms, diarrhea and bloating were significantly more often presented in patients who were admitted to the hospital compared to those discharged home (40% for diarrhea and 14% for bloating vs 18% and 2.6% respectively). Other symptoms such as abdominal pain, heart burn, nausea, vomitus, anorexia, and constipation had not been differently presented in these groups in the mean of statistical significance. C-reactive protein has been also significantly higher in hospitalized group. In case of alanin transaminase (ALT), aspartate transaminase (AST) and bilirubin as markers of possible liver damage only AST (figure 1) has been significantly higher in the hospitalized group. This difference is substantial. There is no statistically significant difference in the levels of ALT (figure 2) and Bilirubin.

**Figure 1.**
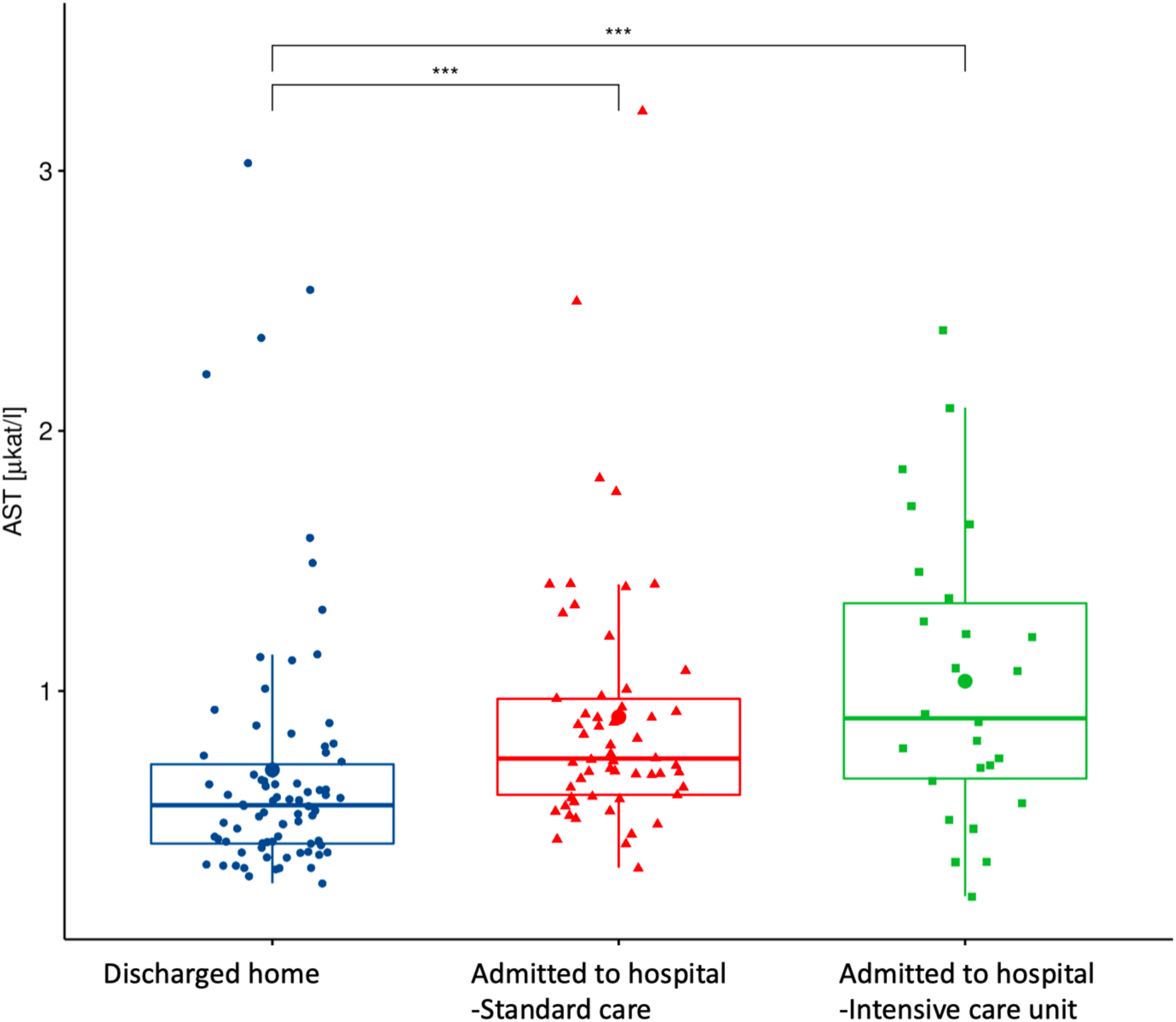
AST: aspartate transaminase; p<0.0001

**Figure 2.**
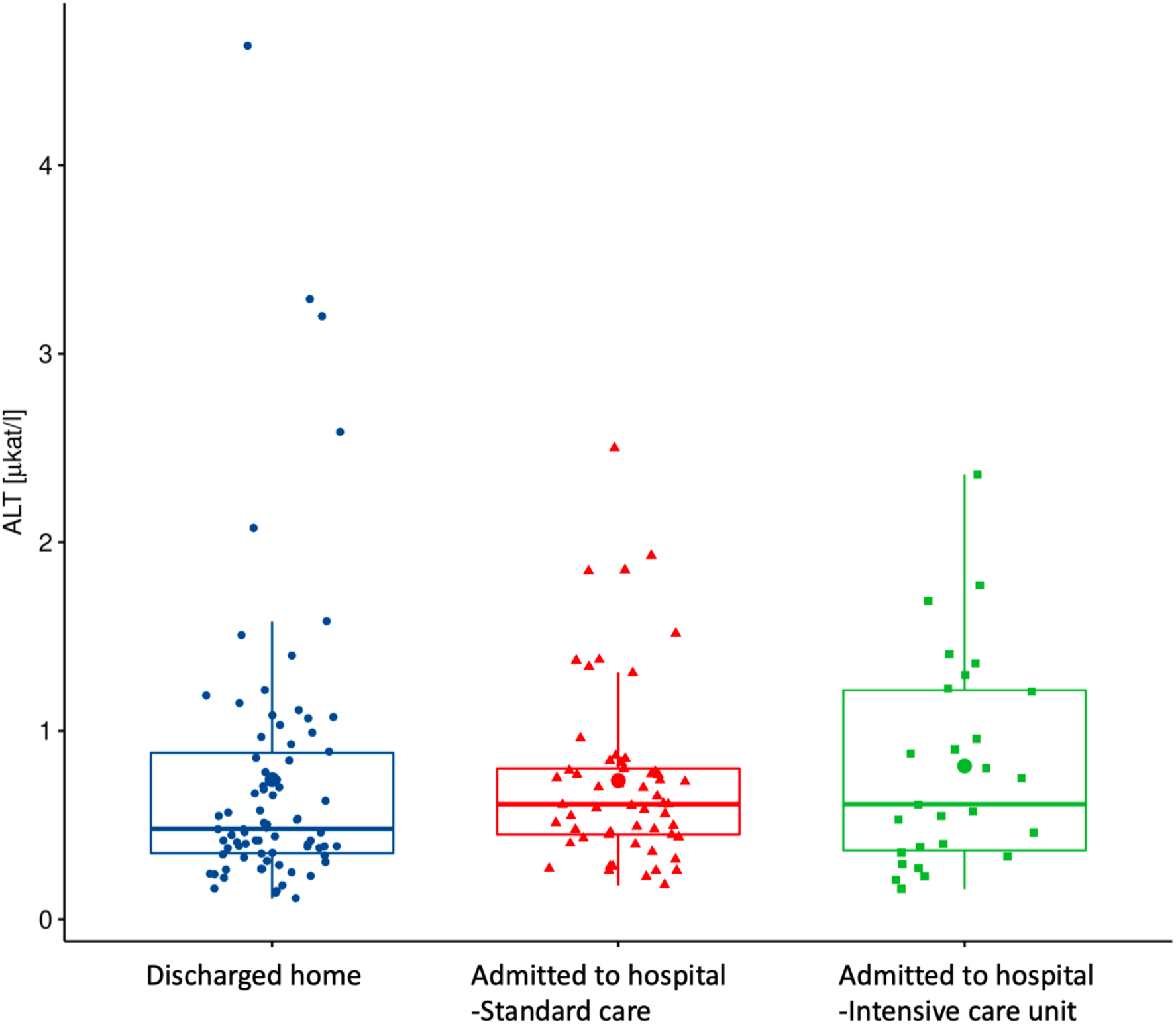
ALT: alanine transaminase; p=0.25

### Predictors of hospitalization based on machine learning

Based on random forest algorithm with the data on demographic characteristics, symptoms and gastrointestinal related laboratory findings in hospitalized and discharged patients, several predictors for risk of hospitalization have been identified. AST has been identified as the most important predictor followed by age or diabetes mellitus. Diarrhea and bloating have also positive importance although much lower than AST. Gastrointestinal symptoms such as nausea, abdominal pain or anorexia have none or negative predictive importance. ROC curve for combined factors is shown at the figure 3 with AUC 0.76.

**Figure 3.**
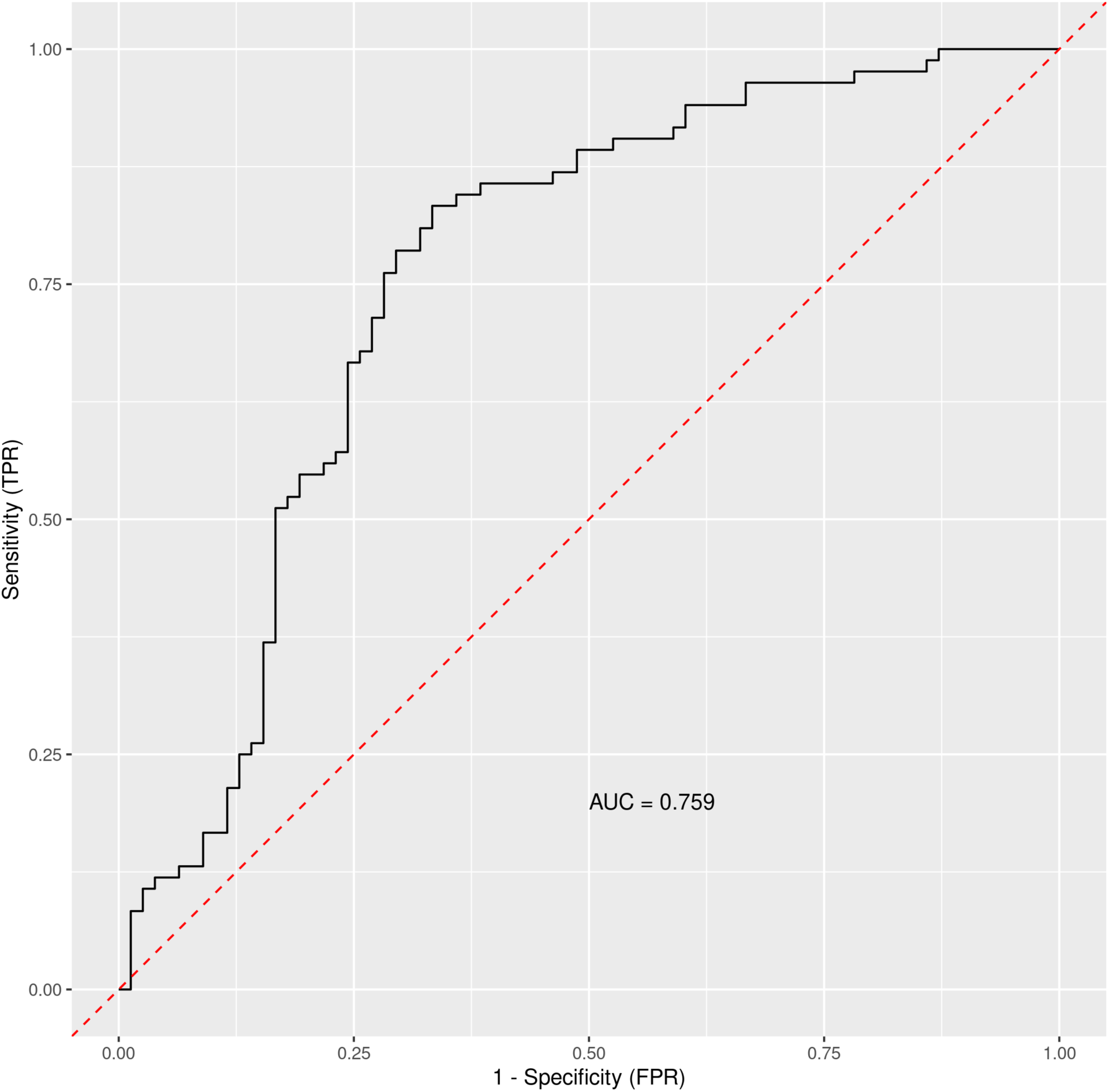
The ROC curve for general COVID-19 symptoms, gastrointestinal symptoms, age, sex, lasting of the symptoms and comorbidities (Diabetes Mellitus, Arterial hypertension and Chronical liver diseases)

When using only liver enzymes (AST, ALT), gastrointestinal symptoms (diarrhea and bloating), chronic liver disease, age and Diabetes Mellitus, the ROC curve (figure 4) for this combination of factors attained AUC 0.799 with AST as the strongest predictor for hospitalization (table 3).

**Table 3.**

|  | **Variable importance** |  |
| --- | --- | --- |
|  | Discharged home | Admitted to hospital |
| **AST** | 0.5729 | 0.2217 |
| **Diabetes Mellitus** | 0.1080 | 0.0288 |
| **Chronic liver disease** | 0.0882 | 0.0061 |
| **ALT** | 0.1025 | -0.0379 |
| **Diarrhea** | 0.0087 | 0.0272 |
| **Age** | 0.0561 | 0.0197 |
| **Bloating** | 0.0052 | 0.0006 |

AST: aspartate transaminase; ALT: alanine transaminase

**Figure 4.**
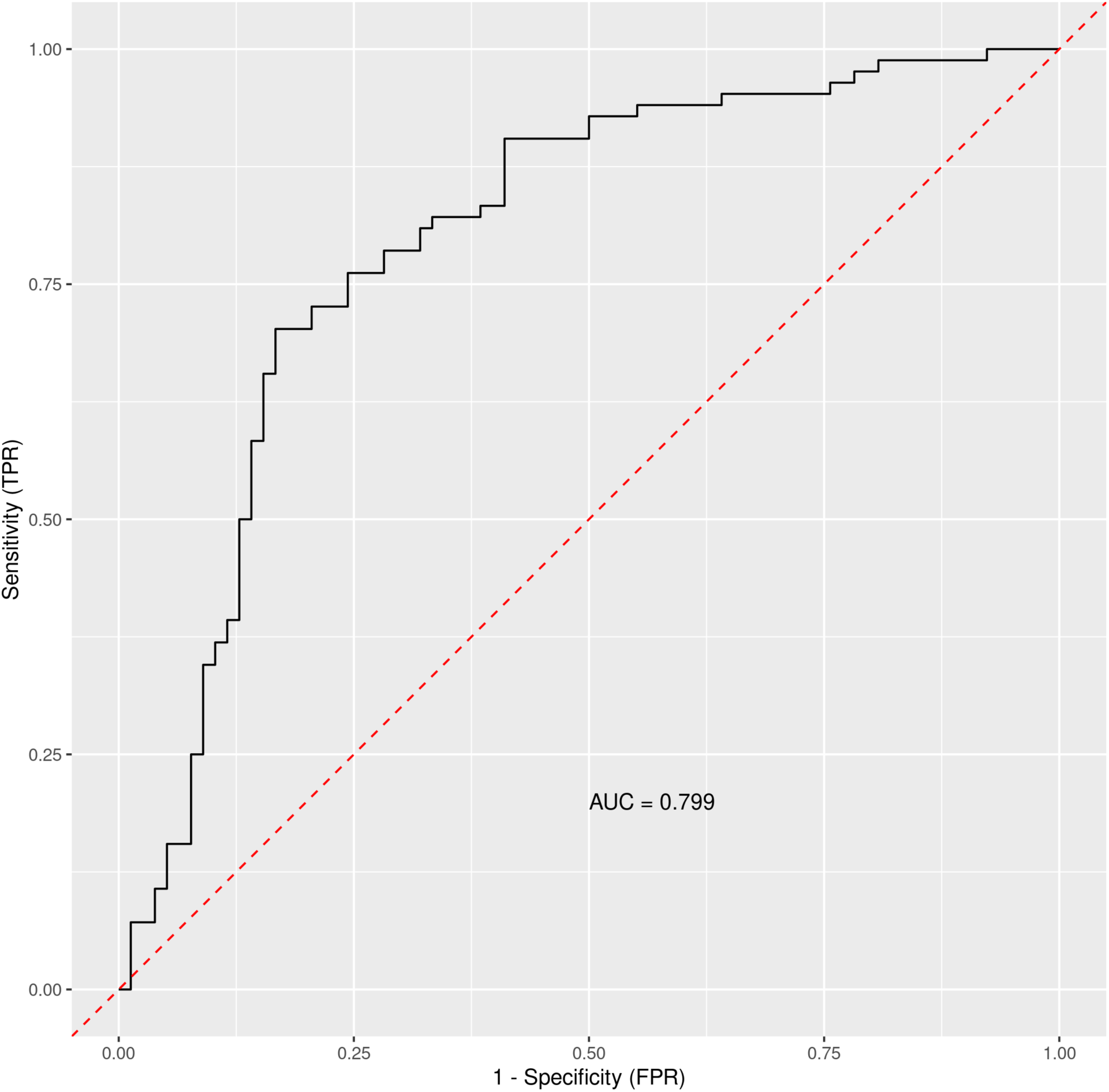
The ROC curve for liver enzymes (AST, ALT), gastrointestinal symptoms (diarrhea and bloating), chronic liver disease, age and Diabetes Mellitus

### Principal components visualization of data

Principal component analysis has been used to get a two-dimensional visualization of the data, with for patients discharged home after ED examination and patients admitted to hospital. Data used for the analysis consist of the data from table 2, that means combination of general patient characteristics, typical COVID-19 symptoms and gastrointestinal symptoms and liver related laboratory results. The PCA plot (figure 5) is showing two distinct clusters which are partially overlapping with tendencies to shift apart.

**Figure 5.**
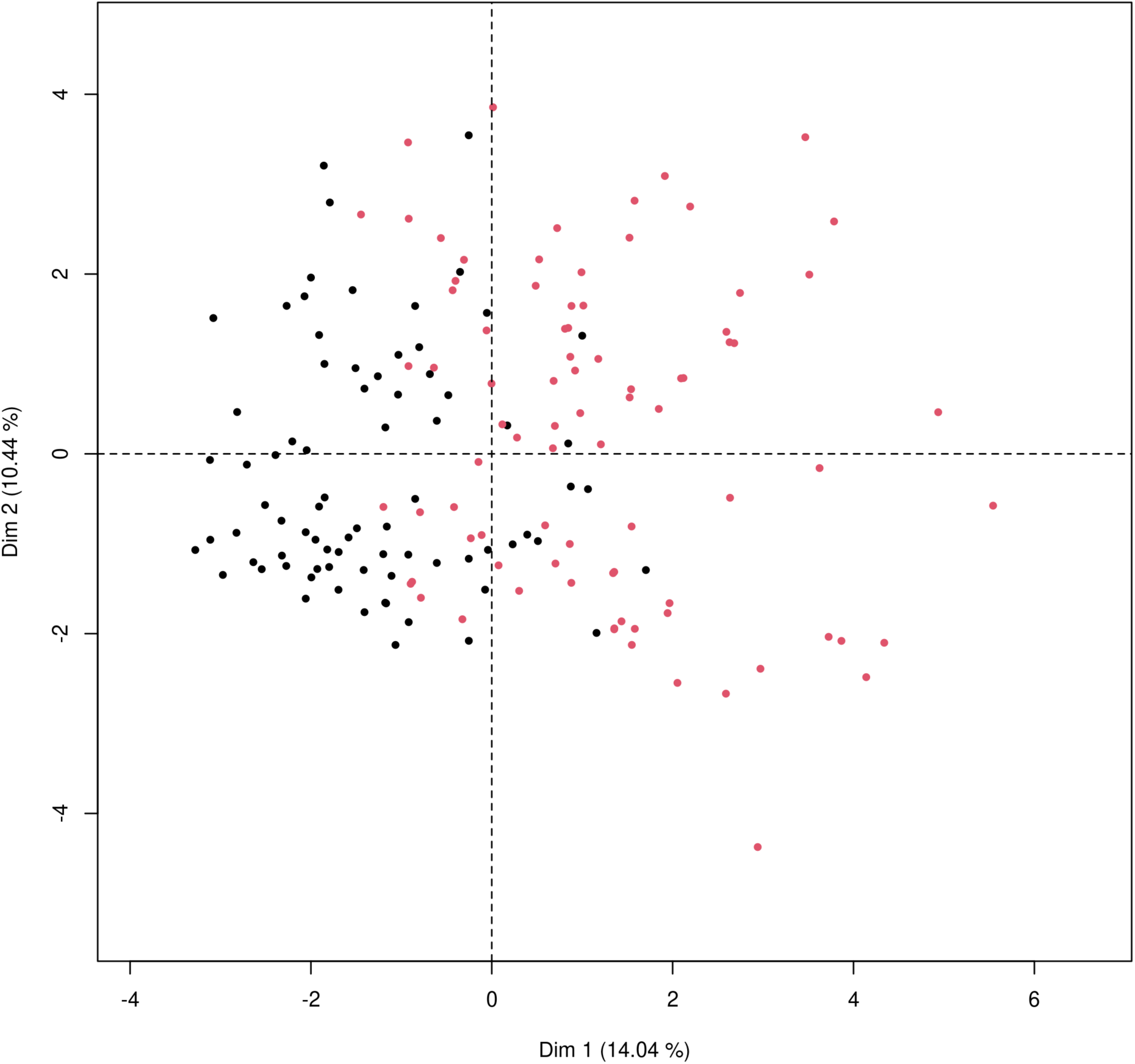
Principal component analysis for mixed type of data to obtain two-dimensional representation of the data on patients who were discharged home (black dots) and those who were admitted to the hospital (red dots)

## Discussion

Several studies and metaanalyses have pointed out the gastrointestinal involvement in the SARS-CoV-2 infection [3], [4], [6], [8], [25]–[27]. The data from the pooled prevalence of gastrointestinal symptoms are varying significantly from 10.5% to 53% between studies [3], [4], [25], [28]. Based on comprehensive metaanalysis by Sultan et al. [4] the pooled prevalence of diarrhea is 7.7%, nausea and vomiting 7.8% and abdominal pain 3.6%. In the presented study we have focused on the presence of diarrhea, constipation, bloating, nausea, heart burn and abdominal pain. Statistically significant differences have been found in the case of diarrhea and nausea when comparing SARS-CoV-2 negative and positive patients. In the group of hospitalized patients (with standard care), the diarrhea was presented in 40% patients and nausea in 21% which is higher compared to some metaanalysis mentioned, but consistent with data considering general presence of gastrointestinal symptoms and gut involvement. When comparing only emergency department group, the presence of bloating is significantly higher in the hospitalized group than those who were discharged home. Interestingly, bloating has lower prevalence in the group of ICU patients than those with standard care management. This could be explained by high subjectivity and interpersonal differences when reporting symptom such as bloating. Considering differences between these two groups of patients, those with more severe course of disease give lower importance to a less annoying symptoms such as heart burn, bloating and nausea when compared to more presented symptoms such as diarrhea, abdominal pain or vomitus. Focusing on the liver enzymes as markers of possible liver impairment resulting from SARS-CoV-2 infection the AST, ALT and bilirubin have been considered for evaluation. The results are showing that median level of liver enzymes has not been elevated in the discharged group. Bilirubin and ALT were also within normal range in the hospitalized group with no statistically significant differences between these two groups. Only AST was elevated over the upper level of the norm in the hospitalized group with progressively higher values in patients who required ICU. The differences between hospitalized and discharged patients are substantially significant. Several previously published data have shown elevation in both transaminases and bilirubin to a different extent ranging from 1% to 53% (mainly ALT and AST accompanied by slightly increased bilirubin concentrations) [3]. In most published data, severe liver alterations were uncommon [29] and the pooled prevalence of liver injury regarding severity was 12% based on the meta-analysis by Mao et al. [3] More severe liver injury was also associated with worse outcomes, including intensive care unit admission and mortality [30].

The pathophysiology of liver involvement in COVID-19 is still not completely understood. The direct viral infection of the liver cells is proposed as a one of causes for liver injury, but the comprehensive studies are scarce. Study with pathological analysis of liver tissues from dead victims of COVID-19 showed no viral inclusions in hepatocytes [31]. Another repeatedly proposed and generally accepted mechanism of liver impairment could be drug toxicity [3]. In order to determine possible influence of recent ATB usage on the elevated AST presented in this paper, two-way analysis of variance (two-way ANOVA) was performed. There are no significant differences between groups with or without recent antibiotics usage. Therefore, we have concluded that ATB usage has no relevant influence on the elevated AST levels. The two-way ANOVA was also performed to assess the relationship between presence of chronical liver disease and AST. There is no statistically relevant difference in AST levels in hospitalized patients with and without chronical liver disease.

Another possible explanation of elevated transaminases could be the result of a systemic inflammation. ALT is an enzyme most commonly found in liver, with small levels in striated muscle tissue and myocardium. On the other hand, AST could be found in liver but also in striated and myocardial muscle, kidneys, brain and red blood cells. AST had been used as a marker for myocardial infarction for a long time before more sensitive markers were identified and implemented to the routine clinical practice [32]. Based on the results of this study and current knowledge of SARS-CoV-2 interaction in human organism it is possible that elevated levels of AST in COVID-19 patients could be result of a systemic inflammation with general tissue hypoperfusion rather than a result of a direct influence of SARS-CoV-2 virus on the hepatocytes or hepatotoxic drug use.

Further we have focused on identifying possible predicting factors for hospitalization in COVID-19 patients using advanced artificial intelligence techniques such as the Random Forest (RF) machine learning algorithm.

Different types of machine learning are being used in increased rate to determine predictors of outcome in various areas of clinical practice from brain trauma injuries [33], radiology [34], oncology [35] to dermatology [36]. Since COVID-19 pandemic is affecting the global population for more than a year a half now and is the cause of immense health crisis in most of the countries in the World new diagnostic tools-machine learning being one of them- and therapeutic methods are rapidly emerging [9]. Shortly after the COVID-19 outbreak various machine learning techniques have been used including taxonomic classification of COVID-19 genomes [10], determining predictors of severe COVID-19 [11] and searching for new potential drug candidates against SARS-CoV-2 viral infection [12]. Another example of successful implementation of artificial intelligence in COVID-19 diagnosis is evaluation of the CT scans detecting SARS-CoV-2 associated pneumonia and differentiate it from community acquired pneumonia and other similar conditions with specificity and sensitivity higher than 90% [13].

So far several studies have been published using random forest machine algorithm for identifying predictors for COVID-19 outcome from wide variety of symptoms, socioeconomical factors [37] and laboratory results with various results [38], [39]. To our current knowledge there are no studies specifically focused on gastrointestinal symptoms and gut related laboratory findings to this date.

In order to assess predictive power of the gastrointestinal parameters and other measured variables for predicting the need for hospitalization the Random Forest Machine Learning algorithm was trained on the data from our study. Results were plotted as the ROC curve obtained from the Out-Of-Bag data. When considering general COVID-19 symptoms, gastrointestinal symptoms, age, sex, lasting of the symptoms and comorbidities (Diabetes Mellitus, Arterial hypertension and Chronical liver diseases) the AUC is 0.76. Measuring the Variable importance, the most important predictor is AST followed by age and diabetes mellitus which are substantially less important. When using only liver enzymes (AST, ALT), gastrointestinal symptoms (diarrhea and bloating), age and presence of chronic liver disease and Diabetes mellitus the AUC is 0.799 with AST as the strongest predictor for hospitalization. Previously published studies which used mostly methods of classical statistic, have identified the presence of gastrointestinal symptoms [40] predominantly diarrhea [41], [42] and elevated liver enzymes [2] as predictors of hospitalization associated with COVID-19. In our data, we have singled out aspartate transaminase (AST) as not only the statistically significantly elevated liver enzyme in patients requiring hospitalization but using artificial intelligence with random forest algorithm AST has proved out to be the most import predictor of hospitalization. Finally, we performed the Principal component analysis for mixed type of data to obtain two-dimensional representation of the data on patients who were discharged home and those who were admitted to the hospital. As could be seen on the plot 5 these two groups are partially overlapping but with clear tendencies to shift apart, which is in accordance with the predictive performance of the studied variables in the random forest algorithm.

## Conclusion

This study has identified elevated AST as the most important predictor for COVID-19 related hospitalizations using machine learning random forest algorithm. We have also shown that SARS-CoV-2 positivity is connected with isolated AST elevation and the level is linked with the severity of the disease. Furthermore, the prevalence of diarrhea and nausea among SARS-CoV-2 positive patients is significantly higher compared to SARS-CoV-2 negative controls. Bloating is occurring significantly more frequently in COVID-19 patients who require hospitalization than those who could be discharged to outpatient care.

## Data Availability

The data that support the findings of this study are available from the corresponding author, upon reasonable request.

## Funding

This publication has been produced with the support of:

The Integrated Infrastructure Operational Program for the project: Research and development of telemedicine solutions to support the fight against pandemic diseases induced Covide-19 and reducing its negative consequences by monitoring the health status of people in order to eliminate the risk of infection in at-risk populations, ITMS: 313011ASY8, co-financed by the European Regional Development Fund.

The Integrated Infrastructure Operational Program for the project: New possibilities for laboratory diagnostics and massive screening of SARS-Cov-2 and identification of mechanisms of virus behavior in human body, ITMS: 313011AUA4, co-financed by the European Regional Development Fund.

By Ministry of Health of the Slovak Republic under the project registration number 2019/44-UKMT-7.

## Authors contribution

P.L. design of the study, drafting the manuscript, data acquisition and analysis, P.B. the concept of the study, R.B. data acquisition and drafting the manuscript, M.P. data acquisition, I.K. data acquisition, I.Z. interpretation of results and drafting the manuscript M.G. statistical analysis and data analysis, P.U. data acquisition, R.H. manuscript control. All authors critically revised the manuscript, approved the final version to be published, and agree to be accountable for all aspects of the work

## Abbreviations

PCR: (polymerase chain reaction)
ALT: (Alanin transaminase)
AST: (aspartate transaminase)
ICU: (intensive care unit)
ED: (emergency department)
ROC: (receiver operating characteristic)
PCA: (Principal component analysis)
RF: (random forest)

